# A systematic review assessing the potential for release of vector species from competition following insecticide-based population suppression of *Anopheles* species in Africa

**DOI:** 10.1101/2021.06.09.21258627

**Authors:** Alima Qureshi, John B. Connolly

## Abstract

**Background:** While insecticide-based vector control can effectively target vector species in areas of high malaria endemicity, such as *Anopheles gambiae* in Africa, residual disease transmission can occur. Understanding the potential role of competitive displacement between vector species could inform both current insecticide-based vector control programmes and the development of future complementary interventions.

**Methods:** A systematic review was conducted to identify studies of insecticide-based vector control of *Anopheles* species in Africa that reported indices for absolute densities of vector species. After screening against inclusion, exclusion and risk of bias criteria, studies were assigned to three categories based on whether they showed population density changes involving decreases in two or more vector species (D), increases in two or more vector species (I), or increases in one vector species concomitant with decreases in another vector species (ID). Category ID studies could thus provide evidence consistent with the release of vector species from competition following the insecticide-based population suppression of *Anopheles* species.

**Results:** Of 5,569 papers identified in searches, 30 were selected for quantitative and qualitative analysis. Nineteen studies were assigned to category D and 1 study to category I. Ten studies categorised as ID provided evidence ranging from weak to persuasive that release from competition could have contributed to changes in species composition. Category ID showed no statistical differences from category D for reductions in malaria transmission and levels of insecticide resistance, but did so for insecticide type, pyrethroids being associated with category ID. A qualitative assessment identified 5 studies that provided the most convincing evidence that release from competition could have contributed to changes in species composition.

**Conclusion:** This review identified evidence that insecticide-based reductions in the density of *Anopheles* species in Africa could facilitate the release of other vector species from competition. While it remains uncertain whether this evidence is representative of most entomological sequelae from the use insecticide-based vector control in the field, 5 studies provided persuasive evidence that insecticide use could lead, at least under some circumstances, to competitive release of non-targeted vector species. These results should inform current and future integrated vector management approaches to malaria control.

## Background

An ecological niche can be considered as the habitat supplying the resources that are required for the survival and reproduction of a species, or the roles and interactions that one species has within a community of other species [1]. Competitive reduction reflects situations where the density of the population of one species decreases, because of competitive interactions, directly or indirectly, with another. Competitive displacement is that more extreme situation where competitive interactions of one species with another causes its local extinction.

Competitive interactions between sympatric vector species could thus potentially play a role in continued disease transmission [1].

Aquatic habitats for mosquito larval development are regarded as having the most impact on adult population densities [2, 3]. Here, competition can occur via (i) exploitation, where individuals compete for limited resources, (ii) interference, where individuals obstruct the development of others, (iii) apparent competition, where there are differential effects of a predator or parasite on co-occurring species, or (iv) oviposition deterrence, where egg-laying by one species is avoided where a competitor species is present [1]. Therefore, differential effects of vector control measures on sympatric vector species could facilitate the release of co-located vector species from competitive displacement or reduction by the targeted species, sometimes termed ‘release from competition’ or ‘competitive release’, leading to ‘niche replacement’ or ‘niche expansion’ of other vector species, potentially facilitating residual or increased transmission of malaria or other vector-borne diseases.

Insecticide-based vector control interventions, such as Long-Lasting Insecticide-treated bednets (LLINs) and Indoor Residual Spraying (IRS), are known to target more effectively vector species that are endophilic, anthropophilic and nocturnal. In Africa, the dominant malaria vectors are *Anopheles gambiae s*.*s*., *Anopheles coluzzii, Anopheles arabiensis* and *Anopheles funestus* [4]. Although *An. funestus* is a complex consisting of 13 species within Africa, of these only the species *An. funestus* is thought to play a role in malaria transmission [5]. While the above four named species typically blood feed nocturnally, *An. funestus* and *An. arabiensis* can also exhibit crepuscular and in some instances diurnal behaviour [6, 7] *An. gambiae s*.*s. and An. coluzzii* are endophilic and anthropophilic in contrast to their sibling species *An. arabiensis*, which displays exophilic, zoophilic behaviour. Thus, sufficient numbers of less-efficiently targeted vector species could possibly remain in the area of vector control and contribute residual, or even resurgent, transmission of malaria.

Differences in characteristics of individual species could further lead to a range of sensitivities of sympatric vectors to insecticides, impacting on behavioural responses seen post-intervention. Some species are known to be behaviourally resilient, exhibiting plasticity, rather than showing evidence of altered innate preferences and subsequent ‘resistance’ to intervention [8]. These differences could potentially aid and further the strength of any competitive interactions seen. Moreover, *An. gambiae s*.*s*. and *An. arabiensis* are sibling species known to co-occur within aquatic habitats [9, 10]. Mesocosm experiments show competition affects development rate of each species in the opposite way. Development time for *An. arabiensis* increases in the presence of *An. gambiae s*.*s*. and decreases for *An. gambiae s*.*s*. in the presence of *An. arabiensis* [11]. In the laboratory, *An. arabiensis* shows reduced survival when reared with *An. gambiae* s.s. compared to when reared alone [12]. When employing targeted vector control measures, it is thus possible to envisage that reduced densities of *An. gambiae s*.*s*. could facilitate competitive release of *An. arabiensis*. However, some caution should be exercised in interpreting whether mesocosm and laboratory-based data are representative of field settings. Indeed, sampling of *Anopheles* larvae from aquatic habitats can yield highly heterogeneous densities [13], making field studies exploring competitive mechanisms potentially challenging.

Nonetheless, understanding the dynamics of changes in vector species composition and any competitive reduction or displacement could be important for the effective use of current insecticide-based vector control programmes and inform the development of novel interventions [14]. While evidence for protective effects of insecticide-based methods of vector control is convincing [15], between 2014-2019 previous global gains in malaria control decelerated, particularly in Africa where the burden of malaria is highest [16]. Although there is extensive literature on studies of competitive interactions between mosquito vector species outside of Africa [1, 17-19], the synthesis of studies within Africa investigating the impacts of insecticide-based vector control on competitive interactions between species of *Anopheles* and other vector species remains more limited [14, 20]. Moreover, while numerous studies from Africa have demonstrated that use of insecticides can result in substantial population suppression of dominant malaria vectors, such as *An. gambiae s*.*s*. or *An. funestus*, many have typically reported changes in the relative proportions, rather than absolute densities, of targeted species to non-target ones [21-24], making it impossible to differentiate reliably between established preferential effects of insecticides on target versus non-target species and any genuine increases in the densities of non-target species that might indicate true competitive release.

Thus, the extent of evidence showing decreases in the densities of targeted vector species and concurrent increases in the density or densities of other species remains unclear. Restriction of a synthesis of the impacts of insecticide-based vector control on competitive interactions to African studies would involve analysis of a more discrete set of vector species than would be the case for a global investigation. This could, therefore, elicit a more precise evaluation of the entomological impacts of insecticide-based malaria vector control programmes in Africa. The aim of this systematic review was thus to critically assess published research to determine whether insecticide-based vector control interventions in Africa targeting *Anopheles* mosquitoes could potentially facilitate the competitive release of other vector species.

## Methods

### Search strategy

A review protocol was developed in accordance with the Preferred Reporting Items for Systematic Reviews and Meta-Analyses [25] guidelines and registered with the PROSPERO International prospective register of systematic reviews (record ID CRD42020194304) [26, 27]. The systematic literature review was undertaken from June to September 2020, using PubMed, Web of Science and Science Direct. Each was searched from inception of each search engine until September 2020. Search terms used included ‘intervention’, ‘insecticide’, ‘insecticide residual spraying’, ‘long lasting insecticidal nets’, ‘anopheles’, ‘vector’, ‘changes’, ‘resurgence’, use of which was based on search strings created from three categories; exposure/treatment, subject and outcome. Boolean operators ‘OR’, ‘AND’, and ‘NOT’ were used to narrow or broaden results. Reference lists of selected articles were also searched. EndNote and Microsoft Excel were used as reference manager software.

### Eligibility criteria

Studies identified were screened against eligibility criteria in specific stages to provide clarity on the causes of exclusion. The first stage was undertaken by AQ and involved screening of title and abstract for the following inclusion criteria for studies: within Africa; species of *Anopheles* mosquitoes included as a mosquito of interest; insecticide-based intervention tested; data contained either before and after, or with and without, intervention arms (Table 1). In the second stage, AQ and JBC conducted full text reviews of all studies based on the previous inclusion criteria plus the following: inclusion of at least two species of mosquito species, with at least one being from the *Anopheles* genus; inclusion of *Anopheles* and other species density outcome data for both before and after or with and without intervention implementation, with accepted density outcome data and trial design criteria, as well as inclusion and exclusion criteria as described in Table 1.

**Table 1.** A summary of inclusion and exclusion criteria used in order to ascertain study eligibility for systematic review.

| **Criteria** | **Inclusion** | **Exclusion** |
| --- | --- | --- |
| **Time period** | Search engine inception (1940) through to September 2020 | - |
| **Language** | English | Any language other than English |
| **Region** | Africa | Any region other than Africa |
| **Trial design** | Randomised control trials; non-randomised control trials; controlled before-and-after studies; cluster randomised trials with distance of more than 2km between different treatment clusters [28]; observation studies; cohort studies; cross sectional studies; small scale trials; interrupted time series studies; studies without a separate control group – provided that the comparator data was sourced from before intervention implementation took place | Laboratory-based studies; opinion pieces; modelling studies; conference abstracts; experimental hut trials; Cluster randomised trials with less than 2km distance between different treatment clusters [29-32] |
| **Intervention type** | Insecticide-based intervention | Any intervention not utilising insecticide |
| **Mosquito type** | At least one *Anopheles* mosquito species, in addition to other mosquito genera | Any mosquito genera combination that does not include *Anopheles*; any non-mosquito species |
| **Mosquito number** | Data from minimum two types of mosquito species required, one of those being of the *Anopheles* genus (see above - 'Mosquito type') | Studies where only data from one mosquito species is recorded |
| **Primary outcome data** | Density measurements including Entomological Inoculation Rate (EIR); Human Biting Rate (HBR); other adult or larval density metrics such as Pyrethrum Spray Catches (PSCs); Human Landing Catches (HLCs); number of mosquitoes per person through human house or catch measured directly through human bait or indirectly through light traps or knockdown catches or baited huts, per stated unit of time, with a maximum time interval measurement of a month/30 days; Rate Ratio based on density data; other proportional data which is calculated using density data | Species composition data without corresponding density data; any other mosquito-based data measurement not sourced from mosquito density data; any study without any density data for the minimum of two mosquito species, one of them being of the *Anopheles* genus; any studies with time interval measurements over a month/30 days |
| **Secondary outcome data** | Proportional data for species identified in primary outcome data; epidemiological supporting data – malaria incidence and infection prevalence in any age group, diagnostically confirmed by microscopy, PCR or diagnostic test; sporozoite rates in mosquitoes; any confounding factors – measures that may have contributed to fluctuations in mosquito density e.g. land use change | Any other outcome data |
| **Data comparator types** | Data from non-intervention arm within a similar or the same locality as intervention arm; Data from an experimental hut set-up with a non-intervention arm as a comparator/control; Data from before intervention roll-out from within the same locality that intervention roll-out took place; Data from intervention arms, when being compared to non-intervention arms, have a high bed-net uptake (>50%); Data to have at least four samples/replicates or measurements per treatment, for all data compared; If not averaged over a year, data comparisons to be sourced from same seasons. If averaged yearly, maximum 30-day intervals acceptable for data from which average is taken | Data with non-intervention and intervention arms not sourced from same/similar locality as intervention arm; Data sourced post intervention roll-out is not sourced from the same or a similar locality from which pre-intervention data was sourced; Data with experimental hut set-up without a non-experimental comparator/control; Data from intervention arms, when being compared to non-intervention arms, have a low bed-net uptake (<50%); Data with less than four samples/replicates per treatment arm for all data compared; Any data comparisons from clearly different seasons (as stipulated in study); Any yearly averages that were not sourced from data taken every 30 days or less |

### Risk of bias

The final stage of eligibility assessment required studies to undergo bias assessment through the *robvis* ROB1 tool [33], and subsequent exclusion of studies that were found to be at risk of ‘High’ bias. Bias and validity assessments were undertaken through a set of outlined bias risks, for each individual study satisfying all inclusion criteria (See Supplementary Table 1 in Supplementary File 1).

### Data extraction

A table was constructed to summarise all study results. AQ extracted data relating to author, year, country, intervention type, insecticide used, vectors involved, replicates for each treatment, percentage mosquito reduction in density (calculated from density data from studies) and corresponding unit of measurement, percentage proportional reduction mosquito and corresponding unit of measurement, percentage reduction in transmission (calculated from transmission data from studies) and corresponding measurement, bias risk, confounding factors, and any other details or comments. Use of percentage reduction meant that any increases in density or transmission were noted as negative numbers, and any decreases were noted as positive numbers (see Supplementary File 2). The following formula was used to establish percentage reduction from extracted data, where ‘first value’ represents data before or without intervention, and ‘second value’ represents data after or with intervention:

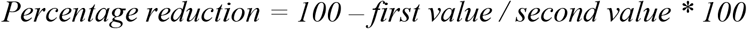

### Categorisation of studies

Following risk of bias assessments, all surviving studies were subsequently assigned to density categories based on reported density changes in vector species:

- Category D - two or more vector species decreasing in absolute densities.
- Category I - two or more vector species increased in absolute densities.
- Category ID - one or more vector species decreased in absolute densities, concomitant with increased density in one or more other vector species.

Individual studies were further stratified by data on location, species, date, trap type, and trap placement (indoors or outdoors), intervention and density. Some studies consisted of multiple density or malaria transmission observations for the stratifications. To differentiate between data arising from these multiple individual observations within studies, from overall data arising from studies, data involving the number of such observations within studies is denoted by the terms ‘observations within studies’ or ‘individual observations’. Multiple observations within a study either fell into the same density category or were heterogeneous for density categories.

### Synthesis and statistical tests

After scrutiny of the quality and quantity of extracted data, a mixed narrative and quantitative approach was adopted for results synthesis. Statistical test application to the dataset was limited due to the heterogeneity of data observations within and between individual studies, such as varieties of vector species identified and multiple sources of mosquito collection (indoors and outdoors, trap collection or HLCs). Considering some studies reported multiple data observations (e.g. data from two different sampling methods or intervention types) any statistical comparisons were made between individual data observations from each study as opposed to whole studies, in order to avoid exclusion of any data. Only studies showing statistically significant density data were included in the review. Statistical tests forming the basis of the meta-analysis consisted of Chi-square tests of independence that considered categorical variables such as intervention type, insecticide use, and insecticide resistance status, and rates of transmission. A Bonferroni adjustment was applied to the result, to minimise the potential for Type I error [34]. Mean transmission intensity change differences between study characteristics such as insecticide resistance, were assessed using a one-way ANOVA.

When extracting and assessing vector density data, to avoid bias, all fluctuations in density that were recorded in each study were considered. For example, if a study took either a “before and after intervention” data collection approach, or a longitudinal data collection approach, or a combination of both, all data satisfying inclusion criteria were extracted and assessed statistically and/or qualitatively.

Where only a proportion of longitudinal data or “before and after intervention” data from within the same study involved category ID, the entire study was assigned to category ID. Transmission data that was tested statistically could be sourced from either sporozoite rates or clinical incidence and prevalence data, with pooled results termed ‘transmission changes’. These data were assessed as individual observations rather than as whole studies, as some studies measured multiple sporozoite rates and therefore had multiple transmission observations for each study.

All statistical testing was carried out using IBM SPSS Version 27. Narrative analysis was carried out in accordance with ‘Synthesis without meta-analysis (SWiM) in systematic reviews: reporting guideline’ [35].

## Results

From 5569 publications screened for this review, 30 studies were eligible for quantitative and qualitative assessments (See Supplementary Tables 2 and 3 in Supplementary File 1). The review and selection process is detailed in Fig 1.

**Fig 1.**
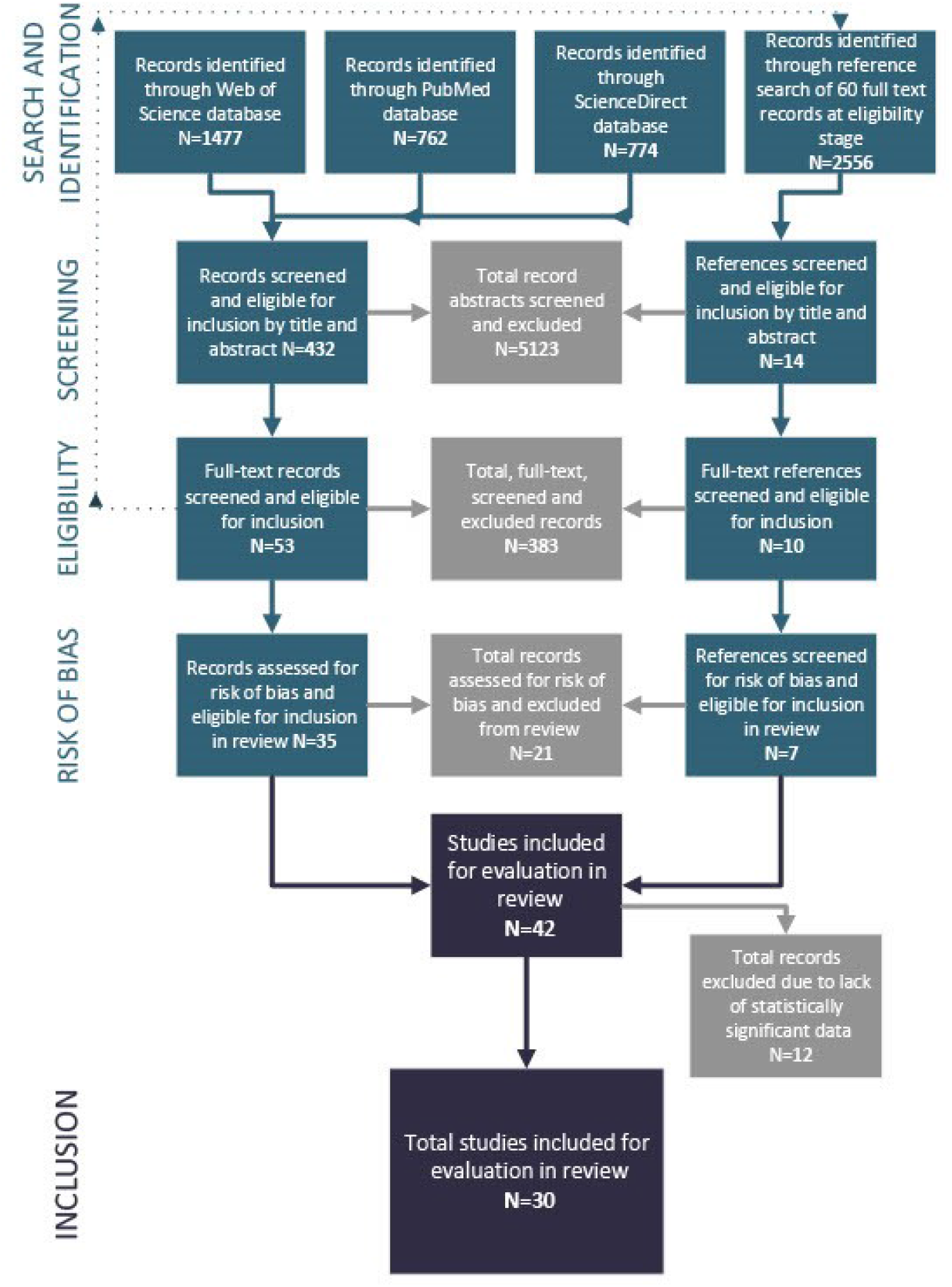
Flowchart summary of all search number results, and exclusion and inclusion numbers at each stage of the search.

### Quantitative analysis

#### Summary of density changes in response to interventions

Of the 30 studies selected for analysis, in total there were 50 and 23 data observations on changes to density data or malaria transmission intensity data (‘transmission data’), respectively. 66% (n=33), 2% (n=1), and 32% (n=16) of individual density data observations within those studies were assigned to category D, I and ID, respectively. Chi square tests of independence were used to compare various characteristics of studies fulfilling the three density change categories. Results are displayed in Table 2. A significant relationship was found between insecticide type used and category, where use of a pyrethroid insecticide was more likely to result in a category ID density change.

**Table 2.**
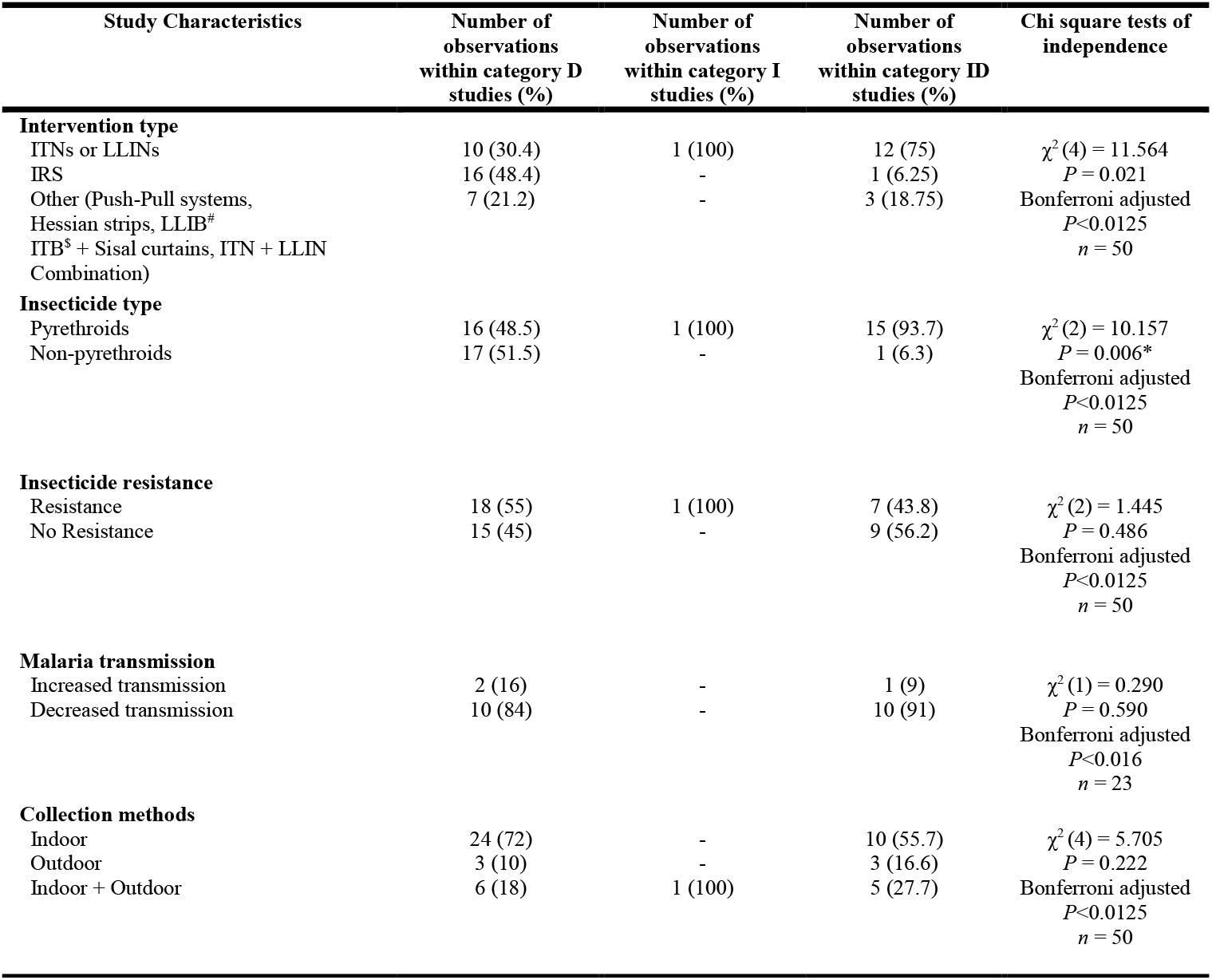
A summary of Chi square comparisons of study characteristics and density change categories. ^#^LLIB: long-lasting insecticide-treated blanket; ^$^ITB: insecticide-treated bednet ^*^Represents a significant difference with Bonferroni correction

#### Insecticide resistance data compared with transmission data

A chi-square test of independence revealed no significant relationship between insecticide resistance status and category of change in malaria transmission (increased, decreased or no change), *X*^2^ (1, *n* = 23) = 0.049, *p* = 0.825. There were no statistically significant differences in mean transmission, between the two insecticide resistance categories (insecticide resistance reported, or not reported) as determined by one-way ANOVA (*F*(1,21) = 17.401, *P* = .945).

#### Transmission data compared to density data

Only a subset of studies that were selected for final analysis recorded transmission data, as this was not a primary observation required for review as per study eligibility criteria. Where category D individual observations within studies had monitored transmission (n=12), average percentage transmission reduction was 48.57, 95% CI [1,96.15]. In category ID (n=11), average percentage transmission reduction was 54.803, 95% CI [30.16,79.44]. There were no statistically significant differences between category mean transmission data as determined by one-way ANOVA (*F*(1,21) = 222.709, *p* = .805).

### Qualitative assessment

Because of the heterogeneity in insecticide-based vector control tools used and vector species studied, it was not considered appropriate to conduct a meta-analysis on density and transmission data on the 10 category ID studies that were identified. Instead, each of these studies were subjected to stringent qualitative assessment, regardless of the outcomes of quantitative analyses.

Qualitative assessment of category ID studies led to the dismissal of 5 studies from further consideration as these were judged to provide only weak evidence for competitive release, such as marginal effects on population densities, or possible alternative explanations that could account for changes in vector species composition such as insecticide resistance or changes in rainfall (see Supplementary File 1). The remaining 5 category ID studies were judged to provide more compelling evidence that release form competition could have contributed to changes in species composition due to release from competition and are described below.

Sougoufara *et al*. [36] examined the impact of the introduction in 2008 of LLINs on densities of the three sympatric species of the *An. gambiae* complex in Dielmo, Senegal. In 2006, prior to LLIN introduction, *An. coluzzii, An. gambiae*, and *An. arabiensis* represented 57%, 19%, and 21% of *Anopheles* captured in human landing catches (HLCs), and accounted for 4.33, 1.45 and 1.62 bites per person per night (bpn), respectively. By 2008, with introduction of LLINs in July, the three species represented 14%, 3% and 83% of proportions of HLCs and had densities of 2.00, 0.44 and 12.12 bpn. Changes in rainfall alone could not account for this change in species densities, as precipitation in 2008 was 834.4mm and 583.4mm in 2006. Indeed, the heavier rainfall observed in 2008 should have favoured higher densities of *An. coluzzii* and *An. gambiae s*.*s*. as both species are typically most abundant in wet season in Dielmo [36].

Over this same timeframe, the prevalence of malaria in children under 14 years of age declined from 36.8% to 12.3%, and from 27.6% to 9.0% for those aged over 15 years, presumably reflecting the shift in composition of the complex to the less endophilic, anthropophilic *An. arabiensis*. By 2010, there was a rebound in the numbers of *An. gambiae*, but not of *An. coluzzii*, which continued to show a decline in density. Although the absolute density of *An. arabiensis* was lower in 2010 than in 2008, it remained the most abundant species, with *An. coluzzii, An. gambiae*, and *An. arabiensis* accounting for 8, 31 and 60%, respectively, of HLC samples. This rebound of An. *gambiae s*.*s*. may have been as a result of the emergence of insecticide resistance, as increases in pyrethroid resistance had been reported in the area in *An. gambiae s*.*l*. [37], although individual species of the complex were not specifically investigated. Adherence to bednet usage may also have contributed to this rebound, as self-reported LLIN usage in Dielmo reduced from 75.6% in 2008 [37] to 58.3% in 2010 [36].

Russell *et al*. [38] also reported a shift in the composition of the *An. gambiae* complex from *An. gambiae s*.*s*. to *An. arabiensis*. In that study, the impact of the 2006 introduction of insecticide-treated nets (ITNs) in rural Tanzania [22] was examined between 2004 and 2009 [38]. Using the absolute density data of the *An. gambiae s*.*l*. reported in Russell *et al*. [38] for 2004 and 2009, and relative density data for *An. gambiae s*.*s*. and *An. arabiensis* reported in the intervention area for 2004 [39, 40] and 2009 [38], the changes in absolute densities of *An. gambiae s*.*s*. and *An. arabiensis* over this period was calculated. These data confirmed an almost complete reversal in the absolute densities of *An. gambiae s*.*s*. and *An. Arabiensis* between 2004 and 2009 for this Tanzanian study (Table 3). Moreover, the authors showed that this shift in species composition was not related to rainfall patterns in the area.

**Table 3.** Evidence from Russell *et al*. [38] for increases in the absolute population densities of *An. arabiensis* concomitant with decreases in absolute densities of *An. gambiae* following rollout of LLINs in 2006 [22]. #Data from Scholte [39] and Scholte *et al*. [40].

| Measurement | Species | 2004 | 2009 |
| --- | --- | --- | --- |
| Absolute density ± standard error (bpn) | *An. gambiae s.l.* | $0.660 \pm 0.006$ | $0.575 \pm 0.008$ |
| Relative proportions ± standard error (%) | *An. gambiae s.s.* | $94.7 \pm 0.05\#$ | 0.5 |
| | *An. arabiensis* | $5.3 \pm 0.05\#$ | 99.5 |
| Absolute density, calculated using relative proportion (bpn) | *An. gambiae s.s.* | 0.625 | 0.003 |
|  | *An. arabiensis* | 0.035 | 0.572 |

Zhou *et al*. [41] reported similar, although less dramatic, observations concerning an increase in density of *An. arabiensis* following decreased density of *An. gambiae s*.*s*. They examined the entomological impacts of the introduction of ITNs in 2006 in villages in Western Kenya between 2002 and 2010. In Kombewa, where the main vectors were *An. gambiae s*.*l*. and *An. funestus s*.*l*., mosquito density decreased by 90% from 2002 to 2007. The 2003 density of *An. gambiae s*.*l*. was 1.3 females per house per night (fhn), with 1.7% of those samples being *An. arabiensis*. In 2009, *An. gambiae s*.*l*. accounted for 0.23 fhn, 61.7% being *An. arabiensis*.

These data allowed the calculation here of the absolute density of An. *gambiae s*.*s*., which decreased more than 14-fold from 1.278 fhn in 2003 to 0.088 fhn in 2009. Over the same timeframe, there was more than a six-fold increase in the absolute density of *An. arabiensis* from 0.022 to 0.142 fhn. In Iguhu, *An. gambiae s*.*s*. decreased more than 11-fold from 2.178 fhn in 2003 to 0.195 fhn in 2006, whereas *An. arabiensis* doubled in density from 0.022 to 0.045 fhn over the same period.

Bayoh *et al*. [42] also examined the species composition of *An. gambiae s*.*l*. in two regions in western Kenya: Asembo, where ITN ownership was circa 90% from 1999 onwards, and neighbouring Seme where ITN coverage was below 5% in 1999, but gradually increased to circa 30% in 2003 and was greater than 70% by 2007. In these communities, *An. gambiae s*.*s*. and *An. arabiensis* were the two dominant malaria vectors, with *An. gambiae s*.*s*. predominating until 1999. After 1999, the proportion of *An. gambiae s*.*s*. relative to *An. arabiensis* steadily declined, so that by 2009 the latter accounted for circa 99% of indoor PSCs and the former 1%. These changes could not be accounted for by rainfall.

Consistent with this proportional data from adult collections, in 2003, the absolute larval density of *An. gambiae s*.*s*. in Seme, with low ITN coverage, was higher than that of *An. arabiensis* and of that found in Asembo where ITN coverage was high. As ITN coverage increased in Seme between 2003 and 2009, the relative proportion of *An. gambiae s*.*s* to *An. arabiensis* obtained in larval collections declined. Thus, the evidence regarding competitive releases of species from this study was consistent with that of Sougoufara *et al*. [36], Russell *et al*. [38] and Zhou *et al*. [41]

Gillies and Smith [43] reported on the impacts on the *An. funestus* group from the application of IRS using dieldrin in the Kihurio village in the South Pare region of Tanganyika (now Tanzania) in East Africa. Before spraying in November 1955, endophilic, anthropophilic *An. funestus* predominated seasonally in box shelter catches while numbers of exophilic, zoophilic *An. rivulorum* were considerably lower (Fig 2).

**Fig 2.**
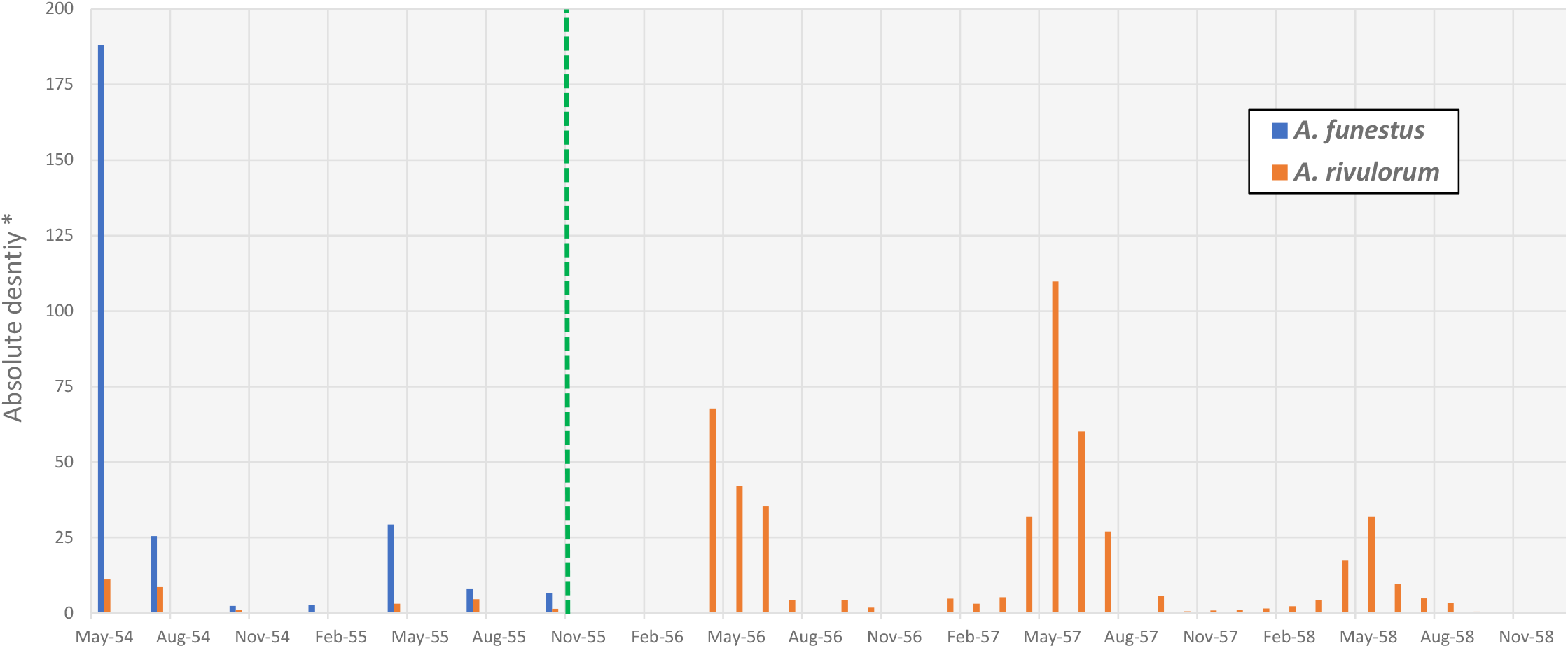
Replacement of *An. funestus* by *An. rivulorum* in Kihurio, South Pare Area, Tanganyika Territory (modern day Tanzania), East Africa, following IRS with dieldrin in November 1955. Data were extracted from Tables I and II and our figure was based on Fig 1 in Gillies and Smith [43]. Green dashed line shows when IRS occured. No samples of *An. funestus* were recorded after January 1956. *Proxies for absolute density were recorded as average monthy catch of mosquitoes from the resting boxes that were used as traps throughout the study area, the number of which varied from day to day. The number of catches per month also varied. Density was therefore calculated using the unit “20 box/days” which represented the average catch of mosquitoes from outdoor traps in the study area on any one day in a month, which allowed comparision between data at different timepoints. Reproduced with permission and adapted from Tables I and II and Fig 1 in Gillies and Smith [43].

After spraying, from January 1956, no sample from box catches was identified as *An. funestus*, while there were substantial increases in the seasonal density of the population of *An. rivulorum*. Although increases in vector densities showed marked seasonal variation (Fig 2), the shift in species composition persisted for more than two years, indicating that such changes in species composition were unlikely to be from environmental or rainfall conditions associated with a particular season. Nor were there any reported changes in irrigation in Kihurio over this time. *An. rivulorum* was identified by the presence of “two spots” on its fifth wing vein, which were never found in *An. funestus*.

The proportion of *An. rivulorum* displaying this phenotype was reported to vary from region to region, so it was possible that the proportion displaying “two spots” changed over the course of the study. However, while this could have exaggerated the magnitude of the effect from the shift in population densities, it would not alone account for the substantial increase in density of *An. rivulorum*.

The authors also reported overlap, albeit incomplete, in the aquatic habitats of *An. rivulorum* and *An. funestus*, found in flooded rice paddies and irrigation channels. *An. funestus*, but not *An. rivulorum*, was also reported to occupy stagnant aquatic habitats. After spraying, the authors did not observe the colonisation of any stagnant water sites by *An. rivulorum*. Rather, they speculated that interspecific competition in overlapping aquatic habitat sites may have accounted for the relative rarity of *An. rivulorum* before spraying. With the near elimination of *An. funestus* following IRS, *An. rivulorum* could have been released from competition, causing such a dramatic expansion in its numbers.

Gillies and Smith [43] also noted that there was a second possibility for release from competition. Other studies in the area had previously reported on the reduction of population density of *An. gambiae*, in addition to *An. funestus* [44]. *An. gambiae* was also found to overlap with *An. rivulorum* in aquatic habitats, so some competitive interactions could be envisaged. However, the annual peak numbers of *An. gambiae* were from January to March, whereas both *An. funestus* and *An. rivulorum* peaked seasonally between April and June. This supports the argument that the increases in density of *An. rivulorum* were caused by the elimination of *An. funestus* leading to the release of *An. rivulorum* from competition in aquatic habitats. Despite its increased population density, *An. rivulorum* did not contribute to malaria transmission in the area because it was principally zoophilic.

## Discussion

The aim of this systematic review was to search for evidence indicating the potential for vector species to be released from competition following insecticide-based vector control of *Anopheles* species in Africa. Of necessity, studies that were included in this review reported population density data for at least two species of mosquito, with at least one belonging to the *Anopheles* genus, both before and after, or with and without, intervention implementation (Table 1).

The stringency of inclusion and exclusion criteria meant that some studies that have previously been cited [1, 20] as providing evidence for release of vector species from competition following insecticide use in Africa, such as Gillies and Furlong [45], were identified but excluded from this analysis because they provided insufficient data on densities of both insecticide-targeted species and other vector species.

Although all category ID studies provided some degree of evidence that opposing changes in species densities were consistent with the release of other vector species from competition (Fig 3 panel A), it is also important consider alternative explanations. Competitive interactions between targeted *Anopheles* species and other vector species could also occur through indirect effects via predator prey interactions where two species could be in apparent asymmetric competition with each other via predator-prey interactions [46], with differential impact of the insecticide on one species leading to the other species to increase in density (Fig 3 panel B). In theory, in such a scenario both species would not need to share habitats, but would need to share predators that could, for example, visit separate larval habitats. Both species would of course still need to co-locate in the same vicinity to be preyed upon by a shared predator.

**Fig 3.**
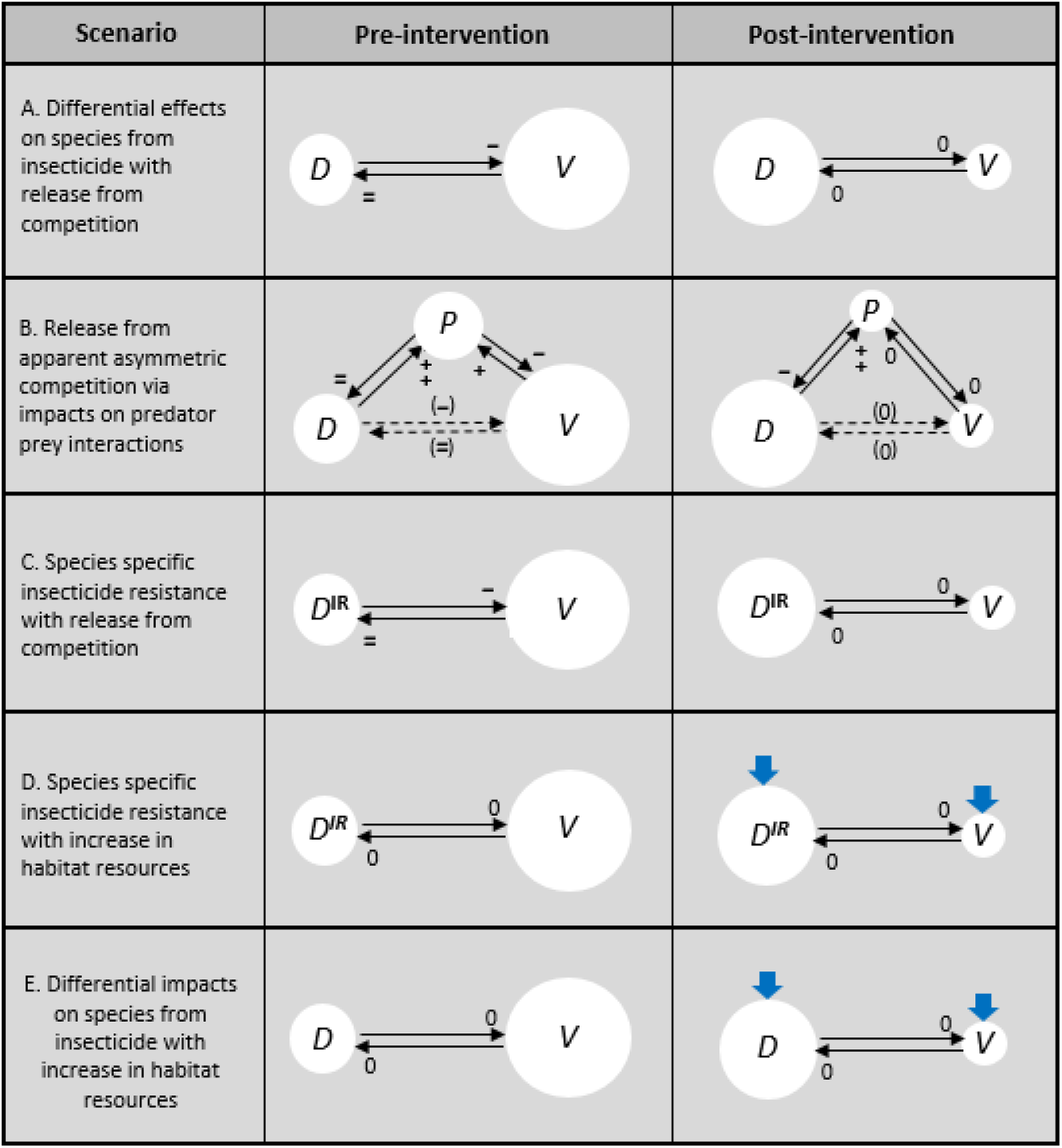
Possible scenarios for changes in densities of species observed in studies from category ID. Intervention involves insecticide-based vector control measure targeting vector species *V* resulting in its decreased population density, concomitant with increased population density of other vector species *D*. White circles represent population densities. Black lines represent completive pressure on a species at the tip of arrowheads, with “+” indicating a positive effect and “+ +” indicating an even stronger positive effect, while “-” indicates a negative effect and “- -” a stronger negative effect. “0” indicates negligible competitive effects. Effects shown in parentheses indicate apparent competitive effects (Holt and Bonsall, 2014). Blue arrows indicate potentially positive effects on population densities from increases in habitat resources, such as increased rainfall. The ^*IR*^ superscript after *D* indicates insecticide resistance in that species. Panels A and B adapted from Connolly *et al*., *Malaria J*. (in press).

Insecticide resistance could also influence changes to species composition. Helinski *et al*. [47] and Zhou *et al*. [48] reported that reduced susceptibility to the insecticide was the most likely cause of divergent changes in species densities resulting from insecticide use. As a result, these category ID studies were excluded from the qualitative assessment. However, even if a species were fully resistant to an insecticide, its density should only be maintained in the presence of the insecticide, but not increase, unless additional factors are also influencing species composition, such as release from competition (Fig 3 panel C), or increases in habitat resources (Fig 3 panel D).In support of this, we found that instances of insecticide resistance were reported for similar proportions of observations across all density categories (55 and 44% in categories D and ID, respectively) and there was no statistically significant relationship between insecticide resistance status and category of density change.

Another possible explanation to account for changes in species composition could involve differential effects of the insecticide on one species over another, combined with an increase in habitat resources, such as increased rainfall or changes to irrigation practices that might augment aquatic habitats. The less affected species could then increase in numbers while the other species reduced in density, without the need to invoke any explanations involving interspecific competition (Fig 3 panel E). Some studies did attempt to address this scenario by searching for evidence for altered patterns of rainfall or irrigation that could support species composition changes, finding none [36, 43], while, one was excluded from the narrative analysis because the reported evidence was judged to be consistent with changes in species composition being driven by changes in rainfall [49].

The most common interventions in category ID studies involved LLINs or ITNs. Species that are endophilic and anthropophagic are behaviourally vulnerable to indoor insecticide-based vector control measures such as IRS and LLINs, which can thus lead to their effective suppression. In the 10 category ID studies identified, there was heterogeneity in species that increased or decreased in density. *An. arabiensis*, a species known to exhibit behavioural plasticity in response to vector control, was found to increase in density in four of these studies but decrease in none. By contrast, *An. funestus*, which shows less behavioural plasticity and has more fixed habitat requirements [50], decreased in density in four of these studies and increased in one. A greater ability to display behavioural evasion of endophilic and anthropophagic needs could thus lead to residual disease transmission. Indeed, mosquito taxa exhibiting such evasive traits may be more accurately described as behaviourally resilient rather than insecticide resistant [8]. Thus, for each species there is likely to be an interplay between physiological status to insecticides and behavioural plasticity that together determine the impact of the intervention.

Limitations also surround the diversity in methods of mosquito collection between each study. Whilst many studies collected indoor in addition to outdoor samples of mosquitoes, some only carried out indoor collections. This may be problematic to interpret, due to the behaviourally selective nature of indoor collections, likely to influence the balance of species densities indoors. For example, *An. gambiae s*.*s*. tends to bite at night indoors and then rest on the walls of dwelling and so is likely to be over-represented in indoor resting catches compared with less endophilic species. However, results indicated that density change type was not influenced by mosquito collection location. Nonetheless, where indoor and outdoor data were reported separately rather than averaged, one study showed that density changes seen from indoor collections (category ID) differed from those seen in outdoor collections (category D) [51]; and two studies showed data consistent with density change type between indoor and outdoor collections [52, 53].

Two additional factors may have contributed to the substantial variation in the strength of evidence supporting release of other vector species from competition found in category ID studies. Firstly, it is know that there can be heterogeneity in observed larval densities in aquatic habitats in the field [13]. This could lead to variability in the propensity for competition between vector species to occur, which could be observed as variability in the frequency and effect size of any releases of other vector species from competition following population suppression of *Anopheles* species. Secondly, release of other vector species from competition could vary temporally. Some releases might only be temporary and occur over relatively short time frames. For example, should LLIN use decline in human behavioural responses to reductions in populations of targeted species, the density of that targeted species could again increase to reduce or reverse any effects of release from competition on the densities of other vector species [36, 37]. Thus, densities reported in in any one study may not have reflected maximal levels of differences in density changes between *Anopheles* species and other disease vectors.

Despite the considerable heterogeneity between category ID studies, potentially involving complex interactions between habitat, climate, environment, and land use changes that may have further contributed to changes in species compositions, qualitative analysis identified five studies reporting significant opposing increases and decreases in vector species that were considered to provide compelling evidence that it was possible for competitive release to occur following insecticide-based population suppression of *Anopheles* vector species [36, 38, 41-43]. In all five cases, highly effective malaria vectors appeared to be replaced with less efficient vectors of malaria transmission. Moreover, in the two of these studies that had also reported on data relevant to disease incidence, indicators of malaria transmission also decreased [41, 54], suggesting that release of vector species from competition did not increase disease transmission under those conditions.

Aquatic habitats for mosquito larval development are believed to have the most impact on adult population numbers [2, 3]. Prerequisites, therefore, for direct interspecies competitive interactions between mosquito species are that they are both sympatric and share the same aquatic habitats. Indeed, it is noteworthy that for the five most compelling studies identified in this review putative competitor species of insecticide-targeted *Anopheles* species were invariably sibling species. Sibling species *An. gambiae s*.*s*. and *An. arabiensis* are known to share larval habitats and competitive interactions between these species have been observed [11], supporting an association of changes in species composition with competitive release [38, 41, 42]. *An. coluzzii* and *An. arabiensis* have also been reported to share aquatic habitats, adding weight to the conclusions of Sougoufara *et al*. [36] that reductions in the density of *An. coluzzii* could have released *An. arabiensis* from competition. Similarly, Gillies and Smith [43] found that *An. rivulorum* replaced it sibling species *An. funestus* following IRS and it was reported that both species shared larval habitats.

Consistent with such observations, this review found no convincing evidence that insecticide-based population suppression of *An. gambiae s*.*l*. could lead to increases in the densities of species outside of the *Anopheles* genus, such as *Ae. aegypti*, a major vector species responsible for the transmission of dengue and yellow fever. Indeed, *Ae. aegypti* and *An. gambiae s*.*l*. do not share aquatic habitats, with the former favouring water tanks, discarded containers, tyres, shells or treeholes [55] and the latter preferring natural aquatic habitats ranging from small, clean, sunlit, ephemeral ones that lack vegetation to larger, riparian, more permanent bodies of water, typically with overhanging vegetation, such as riverbanks with slow-moving water or irrigation channels of rice paddies [56]. Moreover, *Ae. aegypti* is considered an urban dweller whereas *An. gambiae* occupies rural locations.

While it cannot be asserted that the evidence obtained by this review is representative of entomological sequelae from the use insecticide-based vector control in the field, at least 5 studies provided persuasive evidence that insecticide use could, at least under some circumstances, lead to competitive release of non-targeted vector species. These observations should therefore form part of the considerations relating to integrated vector management approaches for malaria control in Africa [57]. More broadly, novel forms of vector control that could in future effectively target malaria vector species [58, 59] might also result in increases in the densities of non-targeted, sympatric vector species that share larval habitats or predators, at least under certain circumstances. Hence, such interventions could effectively reduce the incidence of malaria and thus overall disease burden even amid the background of competitive releases of other non-targeted, sympatric vector species.

## Data Availability

All data and material is presented in the manuscript and supplementary material.

## Declarations

## Acknowledgements

We thank Camilla Beech, Austin Burt, Mamadou Coulibaly, Silke Fuchs, Charles Godfray, Talya Hackett, Andrew McKemey, John Mumford, Samantha O’Loughlin, Seynabou Sougoufara, Frederic Tripet, Geoff Turner, and Katie Willis for useful input over the course of this study.

## Authors’ contributions

AQ and JBC developed the study design and wrote the manuscript. AQ performed the literature searches and conducted screening of abstracts. Both AQ and JBC screened full text articles and assessed selected articles for risk of bias. AQ and JBC read, summarised, and categorised all literature selected from searches. AQ and JBC generated the figures and tables. Both authors read and approved the final manuscript.

### Abbreviations

bmn: bites per man per night
bpn: bites per person per night
fhn: females per house per night
Category D: density of all species decreased with use of insecticide-based vector control
Category I: density of all species increased with use of insecticide-based vector control
Category ID: density of one or more vector species increased while the density of another one or more species decreased with use of insecticide-based vector control
HLC: Human landing catch
IRS: Indoor residual spraying
ITN: insecticide-treated bednet
LLIN: long-lasting insecticide-treated bednet
LTC: light trap collection
PSC: pyrethrum spray catch
UTN: untreated bednet

## Availability of data and materials

Not applicable. All data and material is presented in the manuscript and supplementary material.

## Ethics declarations

### Ethics approval and consent to participate

Not applicable.

### Consent for publication

Not applicable.

