## Supplementary File 1 for "A systematic review assessing the potential for release of vector species from competition following insecticide-based population suppression of *Anopheles* species in Africa"

### A systematic review assessing the potential for release of vector species from competition following insecticide-based population suppression of *Anopheles* species in Africa

Alima Qureshi[1], John B. Connolly[1*]

[1]Department of Life Sciences, Imperial College London, Silwood Park Campus, Ascot, SL5 7PY, UK

#### Risk of bias analysis

A summary of the risk of bias analysis for each study was completed (Supplementary Table 1). N=21 studies categorised as High or Very high for Overall risk of bias were eliminated from this investigation, leaving N=42 studies for further analyses. The most common risk of bias was from 'Confounding factors', where 'Some concerns' were found in N=30 studies. Factors here included any changes in the environment that might have affected the density of vectors other than the insecticide-based intervention itself, such as (1) changes in land use or farming practices such as irrigation in the vicinity of the study, (2) low bed net use (<60%) amongst study populations, in keeping with the Abuja Declaration from the African Summit on Roll Back Malaria [1], or (3) the presence or emergence of insecticide resistance. However, most of these studies (N=28) were assessed as 'Low' or 'Some concerns' for 'Overall' risk of bias, and were therefore included in subsequent analyses whilst taking any potential confounding factors into account. The most common reason (N=19) for studies to be assessed as having a High Overall risk of bias was because they were assigned to High risk of bias in the Missing data or data issues domain. Typically, such studies reported datasets that provided insufficient resolution of information to support unequivocal assessment against our inclusion/exclusion criteria, for example by providing density data graphically, rather than numerically.

30

| Study | Missing data or data issues | Lack of randomisation | Confounding factors | Sample size/ representativeness | Overall |
|---|---|---|---|---|---|
| Abong'o *et al* [2] | Low | Low | Low | Low | Low |
| Akono *et al.* [3] | Low | Low | Some concerns | Low | Low |
| Alegana *et al.* [4] | High | Low | Some concerns | Low | High |
| Antonio-Nkondji *et al.* [5] | Low | Some concerns | Some concerns | Low | Some concerns |
| Bamou *et al.* [6] | Some concerns | Low | Some concerns | Low | Some concerns |
| Bayoh *et al.* [7] | Low | Low | Low | Low | Low |
| Bekele *et al.* [8] | Some concerns | Low | Some concerns | Low | Some concerns |
| Bogh *et al.* [9] | Low | Low | Low | Low | Low |
| Bukhari *et al.* [10] | High | Low | Low | Low | High |
| Chanda *et al.* [11] | Low | Some concerns | No information | Low | Low |
| Coleman *et al.* [12] | High | Some concerns | Low | Low | High |
| Dabire *et al.* [13] | High | Low | Low | Low | High |
| Degefa *et al.* [14] | High | Low | Low | Low | High |
| Fontaine *et al.* [15] | Low | Some concerns | Low | Low | Low |
| Futami *et al.* [16] | Low | Low | Some concerns | Low | Low |
| Gillies and Smith [17] | Low | Some concerns | Low | Low | Low |
| Gimnig *et al.* [18] | Low | Low | Low | Low | Low |
| Gimnig *et al.* [19] | High | Low | Low | Low | High |
| Govella *et al.* [20] | Low | No information | Some concerns | Low | Low |
| Helinski *et al.* [21] | Some concerns | Low | Some concerns | Low | Some concerns |
| Kapesa *et al.* [22] | High | Low | Some concerns | Low | High |
| Kweka *et al.* [23] | Some concerns | Low | Some concerns | Low | Some concerns |
| Labbo *et al.* [24] | High | Some concerns | Low | Low | High |
| Lindblade *et al.* [25] | Low | Low | Some concerns | Low | Low |
| Lozano-Fuentes *et al.* [26] | Low | Low | Low | Low | Low |
| Mahande *et al.* [27] | High | Low | Low | Low | High |
| Majori *et al.* [28] | Low | Low | No information | Low | Low |
| Mbogo *et al.* [29] | Low | Low | Low | Some concerns | Low |
| McCann *et al.* [30] | Low | Low | Some concerns | Low | Low |
| Meyers *et al.* [31] | Low | No information | Some concerns | Low | Low |
| Meyrowitsch *et al.* [32] | Low | Low | High | Low | High |
| Mmbando *et al.* [33] | Low | Low | Some concerns | Low | Low |
| Molineaux *et al.* [34] | Low | No information | No information | Low | Low |
| Musiime *et al.* [35] | Low | No information | Some concerns | Low | Some concerns |
| Mutuku *et al.* [36] | High | Low | Low | Low | High |
| Mwangangi *et al.* [37] | High | No information | Some concerns | Low | High |
| Mwangangi *et al.* [38] | High | Low | Low | Low | High |
| Najera *et al.* [39] | High | Low | Low | Low | High |
| Njan Nloga *et al.* [40] | Low | Some concerns | Low | Low | Low |
| Njoroge *et al.* [41] | High | Low | Some concerns | Low | High |
| Odhiambo *et al.* [42] | Some concerns | Low | High | Low | High |
| Oljira *et al.* [43] | High | Low | Low | Low | High |
| Oloo *et al.* [44] | Low | Low | Low | Low | Low |
| Osse *et al.* [45] | Low | Some concerns | Low | Low | Low |
| Ouattara *et al.* [46] | High | Low | Some concerns | Low | High |
| Pant *et al.* [47] | Low | No information | Low | Low | Low |
| Pant *et al. [48]* | Low | No information | Low | Low | Low |
| Pinder *et al.* [49] | Low | Low | Low | Low | Low |
| Poche *et al.* [50] | Low | Low | Some concerns | Low | Low |
| Protopopoff *et al.* [51] | Low | Low | Low | Low | Low |
| Ratovonjato *et al.* [52] | High | Some concerns | Some concerns | Low | High |
| Russell *et al.* [53] | Low | Low | Low | Low | Low |
| Russell *et al.* [54] | Low | Low | Some concerns | Low | Low |
| Sharp *et al.* [55] | Low | No information | Some concerns | Low | Low |
| Smith and Draper [56] | Low | No information | Low | Low | Low |
| Smith [57] | High | Low | Low | High | High |

| | | | | | |
|---|---|---|---|---|---|
| Sougoufara *et al.* [58] | Some concerns | No information | Some concerns | Low | Some concerns |
| Sougoufara *et al.* [59] | Low | Low | Some concerns | Low | Low |
| Trape *et al.* [60] | Low | Low | Some concerns | Low | Low |
| Wragge *et al.* [61] | High | Low | Some concerns | Low | High |
| Zhou *et al.* [62] | Low | Low | Low | Low | Low |
| Zhou *et al.* [63] | Some concerns | Low | Some concerns | Low | Some concerns |
| Zhou *et al.* [64] | Low | Low | Some concerns | Low | Low |

**Supplementary Table 1**. A summary of the results of risk of bias assessments undertaken on each individual study. Studies were assessed for risk of bias based on four domains, (1) 'Missing data or data issues', (2) 'Lack of randomisation', (3) 'Confounding factors' and (4) 'Sample size/representativeness', each of which contributed to an 'Overall' assessment of risk of bias. In order of ascending risk of bias, studies were classified as 'Low', 'Some concerns', 'High', and 'Very high'. Studies where any domain was designated as High were classified as High for Overall risk of bias. Studies with two or more domains assessed as Some concern, but with the rest assigned as Low, were classified as Some concern for Overall risk of bias. Studies with three domains designated as Low, with the fourth domain assigned to Some concern, were classified as Low for Overall risk of bias.

42

| Study | Category of density change | Mosquito population density decreased | Mosquito population density increased | Mosquito density statistics | Malaria transmission | Malaria transmission statistics |
|---|---|---|---|---|---|---|
| **Akono *et al*. [3]** | I | *n/a* | *An. coluzzi*, Cx. quinquefasciatus** | The biting rates were significantly higher after ITN distribution (*P* = 0.04, *P* = 0.03 respectively). | Increase | EIR difference was 0.83 infective bites per person per night (ib/p/n) and 3.06 ib/p/n before and after LLIN introduction, however the difference between the two periods was not significant (*P* = 0.05). |
| **Antonio Nkondji *et al.* [5]** | D | *An. gambiae*, An. funestus*, An. nili*, An. moucheti** | *n/a* | All species reduced in individual densities. Parity rates were significantly reduced ($\chi^2$ = 55,0; df = 1; P < 0.0001). No other statistical analysis on densities provided. | Decrease* | The average infection rate of malaria vectors significantly decreased from 5.3% to 1.8% after bed net coverage (p < 0.0001). |
| **Bayoh *et al.* [7]** | ID | *An. gambiae* s.s.* | *An. arabiensis** | Density increased significantly with transect sampling position from Asembo to Seme in Poisson regression, and was observed for both sampling events (1st transect sample: risk ratio = 1.28, 95% confidence interval = 1.04-1.58, *P* = 0.006; 2nd transect sample: risk ratio = 1.12, 95% confidence interval = 1.02-1.24, *P* = 0.028) [7]. In the first transect sample, the (density based) proportion of *An. gambiae* s.s. larvae relative to *An. arabiensis* was 16.7% in Asembo but 59% in Seme. In the second transect sample, the proportion of *An. gambiae* s.s. relative to *An. arabiensis* was 9.0% in Asembo and 56.6% in Seme [7].<br>The probability that an individual *An. gambiae* s.l. was identified as *An. gambiae* s.s. increased significantly (logistic regression) with transect sampling position from Asembo to Seme (1st transect: risk ratio = 1.36, 95% confidence interval = 1.09 – 1.68, *P* = 0.006; 2nd transect: risk ratio = 1.77, 95% | n/a | n/a |

| | | | | | | |
|---|---|---|---|---|---|---|
| | | | | confidence interval = 1.49-2.10, *P* < 0.001) [7]. | | |
| **Bogh *et al.* [9]** | D | *An. gambiae s.l.**, *Cx. quinquefasciatus**, *An. funestus** | *n/a* | Comparison before and after intervention showed significant reduction in all *Anophelines* (*P* < 0.05) A decrease in HBI reported for all species (*P* < 0.05). | n/a | n/a |
| **Futami *et al.* [16]** | D | *An. gambiae s.l.**, *An. gambiae s.s.**, *An. arabiensis** | *n/a* | *An. gambiae s.l.* and *An. gambiae s.s.* reduced significantly by year ($\chi^2$ = 940,78; df = 2, P < 0.001). *An. arabiensis* also reduced significantly (P < 0.001). | n/a | n/a |
| **Gillies and Smith [17]** | ID | *An. funestus** | *An. rivulorum** | There was a statistically significant difference between mosquito densities as determined by one-way ANOVA (*F*(3,70) = 5.057, *p* = 0.003. A LSD post hoc comparison revealed that the mean number of *An. funestus* collected before intervention was significantly greater than that of *An. rivulorum* (*p* = 0.017), and that the mean number of *An funestus* collected after intervention was significantly lesser in number compared with *An. rivulorum* (*p* = 0.013). | n/a | n/a |
| **Gimnig *et al.* [18]** | D | *An. gambiae s.l.**, *An. funestus s.l.** | *n/a* | Indoor resting densities of *An. gambiae s.l.* and *An. funestus s.l.* were significantly lower in the intervention houses (58.5%; *P* = 0.010 and 94.5%; *P* = 0.001). | Decrease* | Sporozoite infection rate significantly reduced in *An. gambiae s.l.* in intervention areas EIR estimated to be reduced by 90%. |
| **Govella *et al.* [20]** | D | *An. gambiae s.l**, *Cx. spp** | *n/a* | *An. gambiae s.l.* GLMM analysis (ρ = 1 - Relative Rate (RR) 95% CI = 99.2 [93.89-99%]) *Cx. spp* GLMM analysis (ρ = 1 - (RR) 95% CI = 91.4 [91-92%]). | n/a | n/a |
| **Helinski *et al.* [21]** | ID | *An. funestus** | *An. gambiae s.l.** | Average number of *An. gambiae s.l.* and *An. funestus s.l.* per house per night collected by light traps, significance taken from error bars on graph. | Decrease* | Vector infectivity was 3.2% at baseline and 1.8% three years post-distribution (*p* = 0.001). |
| **Kweka *et al.* [23]** | D | *An. gambiae s.l.**, *Cx.* spp.* | *n/a* | A reduction in mean mosquito densities, data taken from graphs, significance denoted by error bars. | n/a | n/a |

| | | | | | | |
|---|---|---|---|---|---|---|
| **Lindblade *et al.* [25]** | D | *Anopheles*, An. arabiensis*, An. funestus*, Culicines** | *n/a* | Using proportional density data:<br>The proportion of *An. arabiensis* differed significantly between treatment arm (48.8%) and non-treatment arm (22.6%), $P < 0.0001$)<br>*An. funestus* comprised a significantly lower percentage of the total number of *Anophelines* in the intervention arm (11.6%) compared with non-intervention arm (44%; $P = 0.0001$)<br>The indoor resting density of all *Anopheles* was significantly lower in the intervention arm ($P = 0.001$)<br>The number of *Culicines* per house was significantly lower in the intervention arm ($P = 0.001$). | Decrease* | Intervention zone had a lower percentage of anophelines with sporozoites than the non-intervention zone ($P < 0.001$). |
| **Mbogo *et al.* [29]** | D | *An. gambiae s.l.*, An. funestus** | *n/a* | Reduction in ranks of mean zonal densities between before and after intervention for both species (Mann-Whitney U test, $P = 0.0001$). | Decrease | Decrease was not significant |
| **McCann *et al.* [30]** | D | *An. gambiae s.s*, An arabiensis** | *n/a* | Houses with an LLIN had more female *An. gambiae s.s.* and *An. arabiensis* than houses where some people (rate ratios, 95% CI 0.87,0.85-0.89; 0.84, 0.82-0.86; 0.38, 0.37-0.40) or everyone used an LLIN (RR, 95% CI 0.49, 0.48-0.50; 0.39, 0.39-0.40; 0.60,0.58-0.61). | n/a | n/a |
| **Meyers *et al.* [31]** | D | *An. gambiae*, An. melas*, An. coluzzii** | *n/a* | Decrease in densities for all mosquitoes across all sites, with *An. gambiae s.l.* noted as significant through abundance ratios. Significance for *An. melas* and *An. coluzzii* noted through error bars on graph. | n/a | n/a |
| **Mmbando *et al.* [33]** *(indoor data observations only)* | ID | *An. gambiae**, species of *Culex** | *An. funestus*, Mansonia spp.* | All P values represent incidence Rate Ratio (IRR) % protection reductions of number of host-seeking mosquitoes attempting to bite volunteers outdoors in local households with push-pull, versus controls:<br>*An. gambiae* complex, 0.26, $P<0.001$<br>*An. funestus* group -0.48, $P<0.05$<br>*Mansonia* spp. -0.1, $P>0.05$ | n/a | n/a |

| | | | | | | |
|---|---|---|---|---|---|---|
| | | | | *Culex* spp. 0.23, P<0.01. | | |
| **Molineaux *et al.* [34]** | D | *An. gambiae s.l.*, An. funestus*, An. pharoensis** | *n/a* | Positive association between prespraying NBC/PSC ratio for *An. gambiae s.l, An. funestus* and *An. pharoensis,* and the residual density under propoxur. | n/a | n/a |
| **Musiime *et al.* [35]** | D | *An. gambiae s.s*, An arabiensis** | *n/a* | Adjusted Incidence Rate Ratio of 0.07 difference between pre and post intervention, in total human biting rate. Proportions are provided for *An. gambiae s.s.* and *An. arabiensis.* | Decrease | EIR reduced from 129 to 0. |
| **Oloo *et al.* [44]** | ID | *An. funestus** | *An. gambiae s.l.** | Mean house densities/vector/month, with a difference between before and after intervention using data from graphs – *An. funestus* showed decrease whereas *An. gambiae* s.l. showed increase. | Decrease | EIR was reduced by 72% in the intervention village. |
| **Osse *et al.* [45]** | D | *An. gambiae*, Mansonia spp*.* | *n/a* | Significant reduction in blood feeding and rate of both species, and parity rate for *An. gambiae.* | Decrease | Reductions of over 70% of EIR, no statistical analysis provided. |
| **Pinder *et al.* [49]** | D | *An. gambiae s.l.*, An. gambiae s.s.*, An. arabiensis*, Cx. spp* | *n/a* | *An. gambiae s.l.* and sibling species varied by year (before and after intervention),and were slightly lower in the intervention arm. | Decrease* | Reduction in sporozoite positive *An. gambiae* s.l., between years (logistic regression P = 0.039). |
| **Poche *et al.* [50]** | D | *An. gambiae s.s*, An. arabiensis*, An. funestus* | *n/a* | Both *An. arabiensis* and *An. gambiae s.s.* reduced significantly 4 weeks post treatment, in at least one site. No statistical analysis for before and after stated for *An. funestus.* | Increase* | *An. gambiae s.s.* increased in sporozoite rate, significantly in site 1. *An. funestus* also increased, but not significantly. |
| **Russell *et al.* [53]** | D | *An. gambiae s.s.*, An. arabiensis** | *n/a* | Bites per person per night reduced significantly amongst *An. gambiae* complex and *An. funestus* (P < 0.05), after introduction of intervention. | Decrease* | 18-fold reduction in EIR, in protected versus unprotected person<br>4.6-fold reduction with high-bed net (with long-lasting insecticide application) coverage versus untreated nets.<br>Sporozoite prevalence reduced significantly amongst *An. gambiae* complex and *An. funestus* (P < 0.05). |
| **Russell *et al.* [54]** | ID | *An. gambiae* s.s.* | *An arabiensis** | The longitudinal shift in sibling species composition (based on densities) towards *An. arabiensis* was statistically associated | n/a | n/a |

| | | | | | | |
|---|---|---|---|---|---|---|
| | | | | with year (β = -1.152, se = 0.038, $P <$ 0.0001), but was not related to rainfall patterns (β = $-3.079 \times 10^{-4}$, se = $2.04410^{-4}$, $P = 0.132$) [54]. | | |
| **Sharp *et al.* [55]** *(carbamate data observations only)* | D | *An. melas*, An. funestus*, An gambiae* | *n/a* | *An. funestus* and *An. melas* showed 'significant reductions', after second round of IRS (with carbamate). No further explanation of analysis given. | Decrease | No transmission index could be calculated, as all mosquitoes were negative for sporozoites after second spray round. |
| **Smith and Draper [56]** | D | *An. gambiae*, An. funestus** | *n/a* | There was a statistically significant difference between mosquito densities as determined by one-way ANOVA ($F(3,30)$ = 5.770, $p = 0.003$. An LSD Post hoc comparison revealed that densities of *An. funestus* in huts before dieldrin was sprayed, were significantly higher than after ($P < 0.01$), and densities of *An. gambiae* in huts before dieldrin was sprayed, were significantly higher than after spraying ($P < 0.01$). | | |
| **Sougoufara *et al.* [58]** | ID | *An. coluzzii*, An. gambiae** | *An. arabiensis** | The number of bites recorded varied significantly according to species (GLM quasi-Poisson family: $\chi^2 = 9.597$, d.f. = 2, $P < 0.01$) and year (GLM quasi-Poisson family: $\chi^2 = 37.725$, d.f. = 2, $P < 0.001$) [58].<br>The species x year interaction was also significant (GLM quasi-Poisson family: $\chi^2 = 45.869$, d.f. = 4, $P < 0.001$) [58]. | A decrease of 36.8% in 2006 to 12.3% in 2008 in children agead 0-14 years, and from 27.6% in 2006 to 9.0% in 2008 in people aged 15 years or older. Incidence of malaria attacks in the community decreased 57-fold between 2000 and 2012. | n/a |
| **Sougoufara *et al.* [59]** | ID | *An. funestus** | *An. gambiae s.l.** | According to the GLM analysis, species, period, site of collection, rain and month all had a significant effect on the number of bites. Species x period x month interaction was significant, i.e. the effect of months differ by period but differently for *An. gambiae s.l.* and *An. funestus s.l.* | Decrease* | The infection rate varied significantly depending on the periods of the study ($x^2$= 10.648, $P = 0.014$). In P1, a bed net user would have received 40.4 infected bites per year. In p4, a bed net user was expected to encounter 3.2 infected bites per year. |
| **Zhou *et al.* [62]** | D | *An. funestus*, An. gambiae** | *n/a* | *An. gambiae* density in intervention valley reduced by 98% (GLM planned comparison, $F_{1,14} = 7.63$, $P = 0.02$)<br>*An. gambiae* density in the uphill area reduced by 51.6% | Decrease* | Parasite prevalence in the intervention valley dropped significantly from 63.6% before intervention to 16.4% after intervention (GLM planned |

| | | | | | | |
|---|---|---|---|---|---|---|
| | | | | *An. funestus* density in the intervention valley reduced by 85.3% (GLM planned comparison $F_{1,14}$ = 9.16, $P$ = 0.01)<br>*An. funestus* density in the uphill area reduced by 69.2%. | | comparison, $F_{1.24}$ = 309.29, $P$ < 0.0001). |
| **Zhou *et al.* [63]** | ID | *An. gambiae** | *An. arabiensis** | Analysis of species composition (based on densities) illustrated that the proportion of *An. arabiensis* rose significantly in Kombewa from 1.7% in 2003 to 61.7% in 2009. ($\chi^2$ = 65.8, d.f. = 1, $P$<0.0001), but decreased significantly to 11.5% in 2010 (compared with 2009, $\chi^2$ = 119.5, d.f. = 1, $P$ = 0.0001) [63].<br>Species composition (based on densities) change in Iguhu was characterised by a significant increase in the proportion of *An. arabiensis* from <1% in 2003 to 18.8% in 2006 (Fisher exact test $P$ = 0.001), then by a gradual declining trend from 2006 – 2010 (9.2%. Fisher exact test $P$<0.05) [63]. | **Kombewa**<br>Increase*<br>**Iguhu**<br>Decrease* | **Kombewa**<br>Parasite prevalence decreased slightly from 2003 to 2006 (average 52.8%, range 37-78%), then declined sharply (average 8.9%, range 0-25%) during the last half of 2006 (Tukey-Kramer HSD test, P<0.0001). Thereafter it gradually increased throughout 2007 (average 30%) and well into 2008, reaching a monthly rate of 49.6% in 2008, about the same prevalence observed before 2006, and significantly higher than 2007 (Tukey-Kramer HSD test, P<0.05).<br>**Iguhu**<br>Decreasing trend in parasite prevalence was observed from 2002 with a notable decline in 2005, however a sharp declining trend occurred after 2006. The monthly parasite prevalence dropped from an average of 33.8% (range from 18-57%) before July 2006 to 7.5% (range from 2-16%) between July 2006 and December 2008 (Tukey-Kramer HSD test of ANOVA with repeated measure, P<0.0001). Monthly mean parasite prevalence was 13.0% in 2009 which significantly exceeded the 2007/2008 level. |
| **Zhou *et al.* [64]** | ID | *An. gambiae s.l.** | *An. funestus** | *An. gambiae s.l.* differs by being significantly lower in 2008 than 2003 in all sites (no further statistical information provided).<br>*An. funestus* has rebounded significantly in 2008 across all sites, compared to 2003 (unequal variance *t*-test, $t$ = 6.10, d.f. = 7, $P$ < 0.001). | Decrease* | Parasite prevalence in school children decreased sharply at all sites from 2003-2008 ($\chi^2$ tests, $P$ < 0.01 at all sites). |

**Supplementary Table 2. Studies with statistically significant changes in absolute densities of vector species.** Statistical differences were sourced from raw data analysis in each of the studies selected for detailed analysis of evidence for release from competition of vectors after suppression of *Anopheles* vector by insecticide-based intervention. *indicates statistically significant results.

| Study | Category of density change | Mosquito population density decreased | Mosquito population density increased | Mosquito density statistics | Malaria transmission | Malaria transmission statistics |
|---|---|---|---|---|---|---|
| **Abong'o *et al.* [2]** | ID | *An. funestus** | *An. arabiensis* | A statistically significant difference-of-differences was observed between period of mosquito collection and intervention status post-IRS indicating a stronger decline of *An. funestus* in the IRS sites compared to the non-IRS sites (RR = 0.06, 95% CI:0.03-0.13, p<0.001).<br>The mean numbers of *An. arabiensis* collected in indoor CDC-LTs in both interventions and non-intervention sites increased in the post-IRS compared to pre-IRS period, with a statistically different increase only for the non-IRS sites (IRS sites: RR = 1.39. 95% CI:0.78-2.47, p = 0.266; non-IRS sites: RR = 3.06. 95% CI 1.59-5.92, p = 0.001).<br>*An. funestus* comprised over 80% of the total *Anopheles* collected in both intervention and non-intervention sites before IRS. While *An. funestus* remained dominant in non-intervention sites after IRS, *An. arabiensis* formed the bulk of all *Anopheles* collected in the intervention sites after IRS. | Sporozoite infection rates in both *An. funestus* and *An. arabiensis* decreased post IRS. | n/a |
| **Bamou *et al.* [6]** | ID | *An. moucheti** | *An. marshalli, An paludism, An. ziemannii* | In Olama, a significant decrease in mosquito densities was recorded (GLM, Wald $\chi^2$ = 16.27, *P* = 0.0001).<br>The increase in density of mosquitoes from which proportions of mosquitoes were determined, was not significant (GLM, Wald $\chi^2$ = 0.18, *P* = 0.66). | A decrease in EIR was recorded in both Olama (92%) and Nyabessan (26%), between 2000 and 2016. | n/a |

| | | | | | | |
|---|---|---|---|---|---|---|
| **Bekele *et al.* [8]** | D | *n/a* | *An. gambiae s.l., An. pharoensis, An. coustani* | No statistical analysis provided | Decrease* | The *Plasmodium* prevalence was decreases in the kebeles that were covered with ITN+IRS as opposed either only ITN or no intervention (P < 0.05). |
| **Chanda *et al.* [11]** *(ITN data observations only)* | D | *An. funestus, An. arabiensis* | | No statistical difference between figures | No vectors tested positive for sporozoites | n/a |
| **Chanda *et al.* [11]** *(IRS data observations only)* | I | *n/a* | *An. gambiae s.l., An. arabiensis* | No statistical difference between figures | No vectors tested positive for sporozoites | n/a |
| **Fontaine *et al.* [15]** | D | *n/a* | *An. funestus, An. gambiae* | *An. funestus* man biting rate reduced from 1.6 b/m/n to 0.004 b/m/n<br>*An. gambiae* 4.6 b/m/n to 0.01b/m/n. | n/a | n/a |
| **Lozano Fuentes *et al.* [26]** | ID | *An. arabiensis* | *An. gambiae s.s.*, An. funestus s.s.* | No statistical tests undertaken<br>Before/After data used from graphs, indicating confidence intervals. Treatment vs. control data not used, due to both areas being within 0.5km of one another. | n/a | n/a |
| **Majori *et al.* [28]** | D | *n/a* | *An. gambiae s.l.*, An. funestus* | *An. gambiae s.l.* was reduced from 137.09±59.64 females/room in the pre-treatment catch to 0 after treatment, this reduction was significant ($F$ = 6.01, $df_1$=2, $df_2$=52. $P$<0.01).<br>A similar reduction is reported for *An. funestus,* though a statistical difference is not mentioned. | n/a | n/a |
| **Mmbando *et al.* [33]** *(outdoor data observations only)* | D | *An. gambiae**, species of *Culex* | *An. funestus, Mansonia spp.* | All P values represent IRR of % protection reductions of number of host-seeking mosquitoes attempting to bite volunteers outdoors in local households with push-pull, versus controls:<br>*An. gambiae* complex 0.49, P<0.005<br>*An. funestus* group -0.48, P>0.05<br>*Mansonia* spp. -3.2, P>0.05<br>*Culex* spp. 0.02, P>0.05. | n/a | n/a |
| **Njan Nloga *et al.* [40]** | ID | *An. moucheti, Mansonia spp.* | *Cx. quinquefasciatus* | 'Slight' increase in *Cx. quinquefasciatus* after eight months of introduction of | Decrease* | Prevalence of malaria parasites was reduced significantly by 40.3% (Z = |

| | | | | | | |
|---|---|---|---|---|---|---|
| | | | | bednets is mentioned, but no statistical analysis performed. | | 4.54), in subjects less than 15 years after the installation of bednets |
| **Pant *et al.* [47]** | ID | *An. gambiae* | *An. funestus* | n/a | n/a | n/a |
| **Pant *et al.* [48]** | D | *n/a* | *An. gambiae, An. funestus* | No statistical testing undertaken | n/a | n/a |
| **Protopopoff *et al.* [51]** | D | *n/a* | *An. gambiae s.s.*, An. arabiensis, Cx. spp.* | No significant reduction for *An arabiensis* or *Culex spp.* But *An. gambiae s.s.* density was 85% lower (adjusted IRR 0.15, 95% CI: 0.05-0.44, p = 0.001), in the IRS+ITN arm compared to ITN only arm. | Decrease | 0.73 OR (95% CI 0.21-2.54), P=0.607 for Sporozoite rate |
| **Sharp *et al.* [55]** *(pyrethroid data observations only)* | ID | *An. melas*, An. funestus** | *An. gambiae* | Numbers taken from longitudinal data, after first round of IRS. No statistical analysis explained in text, apart from *An. melas* and *An. funestus* showed 'significant reductions'. | Decrease | After first spray round, sporozoite prevalence reduced from 6.0%,8.3% and 4.0% for *An. gambiae s.s., An. melas* and *An. funestus* to1.8%, 3.1% and 2.3% respectively. No statistical analysis information provided. |
| **Trape *et al.* [60]** | ID | *An. funestus* | *An. gambiae s.l.* | *An. funestus* mean monthly human biting rate decreased substantially after the introduction of LLINs, but the mean monthly human biting rate for *An. gambiae* increased. Data taken from graph. No statistical analysis available for time points. | Decrease*, and then increase* | Initial decrease between December 2010 and August 2008 (p=0.0001 by two-sided binomial exact test). Increase between January 2007 and July 2010 (p<0.0001 by Fisher exact test). |

**Supplementary Table 3. Studies without statistically significant changes in absolute densities of vector species.** A description of those studies that did not meet statistical significance testing criteria; thus had to be excluded from review

**Category ID studies dismissed from qualitative assessment in this review, as providing only weak evidence for competitive release or suggesting alternative explanations for changes in vector species composition**

***Marginal changes in population densities***

Mmbando *et al.* [33] examined the entomological effects of a push-pull system, where mosquitoes are both repelled from human hosts by dispensers containing transfluthrin and attracted to lethal odour-baited landing boxes in Tanzania. While the authors reported decreases in the densities of *An. gambiae s.l.* and species of *Culex* with concomitant increases in the densities of *An. funestus* and species of *Mansonia*, these differences were relative low, rendering results inconclusive.

Oloo *et al.* [44] examined the impact on population densities of *An. gambiae s.l.* and *An. funestus* between June 1991 and June 1992 following the introduction in May 1991 of permethrin impregnated sisal curtains. They found indoor biting rates for both *An. gambiae s.l.* and *An. funestus* were substantially reduced in the intervention village compared with control sites. However, there was an increase in biting rates for *An. funestus* compared to controls between April and July 1992 that was not observed in *An. gambiae*. However, the duration of the study was too short to provide any data beyond June 1992 after one year of investigation.

***Insecticide resistance***

Helinski *et al.* [21] examined the entomological impacts of LLIN universal coverage in March/April at four sites in midwestern Uganda, of which for Buliisa only, they recorded density data for both *An. gambiae* s.l. and *An. funestus*. In Buliisa, light trap collection (LTC) data revealed that 66% of all samples were *An. funestus*, of which 97% were *An. funestus s.s*, 21% of samples were *An. gambiae s.s.*, and 13% *An. arabiensis*. By May 2011, the density of *An. gambiae s.l.* had increased significantly from pre-intervention levels while *An. funestus* had almost completely disappeared. Unfortunately, the relative proportions of *An. gambiae s.s*. and *An. arabiensis* over these different time points were not reported. *An.*

*funestus* resurged over 2012 so that it again became the dominant species. The authors tested for the *kdr* L1014S mutations in *An. gambiae s.l.* samples and found that its frequency increased from 72% of *An. gambiae s.s.* in 2009 to 87, 96 and 86% in 2010, 2011 and 2012, respectively, although phenotypic resistance was not assessed. By contrast, no samples of *An. arabiensis* were positive for the *kdr* mutation over this period. Thus, the increase in density of *An. gambiae s.l.* in 2011 could have been caused by increases in numbers of insecticide resistant *An. gambiae s.s.*, or by increases in numbers of *An. arabiensis* caused by changes in inter-species competitive interactions, or by a combination of the former and latter. A possible explanation for the resurgence in *An. funestus* was the emergence of insecticide resistance in that species, although neither its *kdr* status nor phenotypic resistance was tested in the study. The rebound of *An. funestus* may also have been caused by the increased density of a species other than *An. funestus s.s* from the group. It is also worth noting that resistance in a species would be expected to maintain the density of a population in the face of insecticide use, but not to yield an increase in density unless there was additionally some release from competition on the resistant species.

Zhou *et al.* [64] investigated the impact of mass distribution of ITNs in western Kenya between 2006 and 2015. The percent of ITN usage increased in all villages from averages of 23.30 ± 1.08, 43.77 ± 8.44, and 85.59 ± 14.01 in 2006, 2010 and 2011, respectively. In 2006, *An. gambiae s.l.* was the predominant vector in Iguhu and Marani, and *An. funestus* in Kombewa. *An. gambiae s.l.* and *An. funestus* decreased in density in all villages between 2006 and 2008, but then increased in 2011 and 2015. *An. gambiae s.l.* became the dominant vector in Kombewa, and *An. funestus* in Marani. In Marani, for both vectors, there were greater densities in the ITN arm of study, compared to non-ITN arm. However, these effects appear to have been mediated by pyrethroid resistance in *An. gambiae* [65, 66], as suggested by the authors for *An. funestus*. Even though these increases in densities occurred parallel to increases in bednet uptake, malaria transmission reduced over the period of the study.

***Rainfall***

Sougoufara *et al.* [59] investigated the impact of the introduction and two rounds of renewal of LLINs in Dielmo, Senegal over four different time points. P1 was during the pre-intervention period between July 2006 and June 2008. P2 was after the first implementation of LLINs between July 2008 and June 2011. P3 was after the first renewal of LLINs between July 2011 and June 2014. P4 was after the second renewal of LLINs between August 2014 and April 2016. The density of *An. funestus* rapidly declined following the initial introduction of LLINs and remained low at 12.5, 1.0, 1-2, and 1-2 bpn in P1, P2, P3, and P4, respectively. By contrast, the density of *An. gambiae s.l.* initially increased following introduction of LLINs but subsequently decreased following the second renewal of LLINs, with 8.7, 12.3, 4.8, and 1.7 bpn in P1, P2, P3, and P4, respectively. Of note, before the intervention, *An. funestus* was the dominant species involved in year-round malaria transmission, whereas the prevalence of *An. gambiae* was associated with the rainy season.

During the pre-intervention period there were two years with lower than normal rainfall which reduced water resources required for larval habitats so that *An. funestus* numbers had already started to reduce during P1 between 2006 and 2008 and continued to decline post-intervention. By contrast, the increase in density of *An. gambiae s.l.* in P2 was associated with increases in rainfall between 2008 and 2010. It was only following renewal of LLINs that *An. gambiae s.l*. declined in P3 between July 2011 and June 2014. Thus, the authors proposed that the increase of *An. gambiae s.l.* could be explained by the increase in rainfall as a paramount resource required for the development of larval habitats and numbers of adult *An. gambiae s.l.*
